# Examining gender differences in opioid, benzodiazepine, and antibiotic prescribing

**DOI:** 10.1101/19003533

**Authors:** Tymor Hamamsy, Suzanne Tamang, Anna Lembke

**Affiliations:** Center for Data Science, New York University, New York, New York, United States of America; Biomedical Data Science, Stanford University School of Medicine, Stanford, California, United States of America; Department of Psychiatry and Behavioral Sciences, Stanford University School of Medicine, Stanford, California, United States of America

## Abstract

While gender differences have been explored across several areas of medicine, our study is the first to present a systematic comparison of drug prescribing behavior of male and female providers, including opioid, benzodiazepine, and antibiotic prescribing. Our work is of particular relevance to the current opioid crisis and other iatrogenic harms related to injudicious prescribing. Our objective is to explore prescribing differences between male and female providers across medical specialties and for different prescription drug categories in Medicare Part D. To this end, we performed a descriptive, retrospective study of 1.13 million medical providers who made drug claims to Medicare Part D in 2016, analyzing by gender, specialty, and drug category. We found that male providers across diverse specialties prescribe significantly more medications, including opioids, benzodiazepines, and antibiotics than female providers by volume, cost, and per patient. These observed gender differences in prescribing, while agnostic to the quality of care provided, nonetheless inform the design of prevention strategies that seek to reduce iatrogenic harms related to prescribing.

## Introduction

Opioid, benzodiazepine, and antibiotic prescribing has been linked to increased morbidity and mortality in the United States. Among the more than 11 million people who misuse prescription opioids, the majority gets them directly or indirectly from a prescription.^1^ Among heroin users, 75% were first exposed to opioids through a prescription.^2^ Benzodiazepine prescriptions have increased over 67% since 1999, and have contributed to an almost eight fold increase in mortality rates due to benzodiazepine overdoses, especially when combined with opioids.^3^ Antibiotic resistance, a result of antibiotic overprescribing, has the potential to affect people at any stage of life, and poses substantial risks to the viability of the healthcare, veterinary, and agriculture industries, making it one of the world’s most urgent population health problems.^4^

We examined differences in opioid, benzodiazepine, antibiotic, and overall prescribing among male and female Medicare Part D prescribers (2016). As we consider tailored interventions to educate prescribers about the problems of overprescribing, physician factors that drive prescribing take on heightened urgency.

## Methods

We analyzed data from Medicare Part D prescribers from 2016 drug claims submitted to the Centers for Medicare and Medicaid Services (CMS). Collectively, there were 40.4 million Medicare Part D patients, and over 1.4 billion Medicare Part D claims, costing over $146 billion. This is a nationally representative sample of Medicare beneficiaries, in which 85% are over 65.

For each of the 1.13 million unique Medicare Part D prescribers, we mapped the 223 clinical specialties to broader, more common names, based on the CMS provider taxonomy code set. We focused on the potential influence of gender on the number of provider’s patients a provider sees, by exploring the total drug, opioid, and antibiotic beneficiaries of doctors, controlling for the provider’s specialty. We examined gender differences in opioid, benzodiazepine, and antibiotic prescribing for each specialty. For each specialty and drug category, we calculated the median number of claims per patient (effectively the drugs prescribed per patient), for male and female prescribers. The Mann-Whitney test was used to determine the significance of the differences between medians, statistically testing if females prescribed less than males using a Bonferroni corrected *p*-value threshold of .001. Instead of mean estimates, we chose to compare median prescribing because it resulted in a more robust statistic for these data (Medicare Part D is full of prescriber outliers, and the majority of them are male). To adjust for the differences in the patient populations of male and female prescribers (e.g., prescribers are more likely to treat a patient of their own gender), we applied linear models to account adjust for the differences in the patient populations, including variables for the gender balance, average risk score, and the average age of a prescriber’s patients. We adjusted for the age of prescribers, creating a variable for how long the provider had been practicing, based on the year in which they graduated from medical school. Although we performed the same analysis for all drugs, opioids, benzodiazepines, and antibiotics, due to the small number of specialties and providers that prescribe benzodiazepines, we exclude benzodiazepines when a large sample size is important (e.g., in linear modeling).

## Results

Female providers accounted for 42% of the 2016 Medicare Part D prescribers, and 33% of the overall claims. Based on the analysis of provider-level prescribing patterns. we observed a consistent trend of male providers prescribing more drugs per patient than female providers, with a few exceptions by specialty.

Although we observed some variation, male providers prescribed drugs for substantially more patients across almost all of the top prescribing specialties. By provider specialty, Figure 1. shows a heatmap that visualizes the percent difference of male and female providers’ median total drug cost, as well as total, opioid, and antibiotic Medicare Part D beneficiaries.

**Figure 1:**
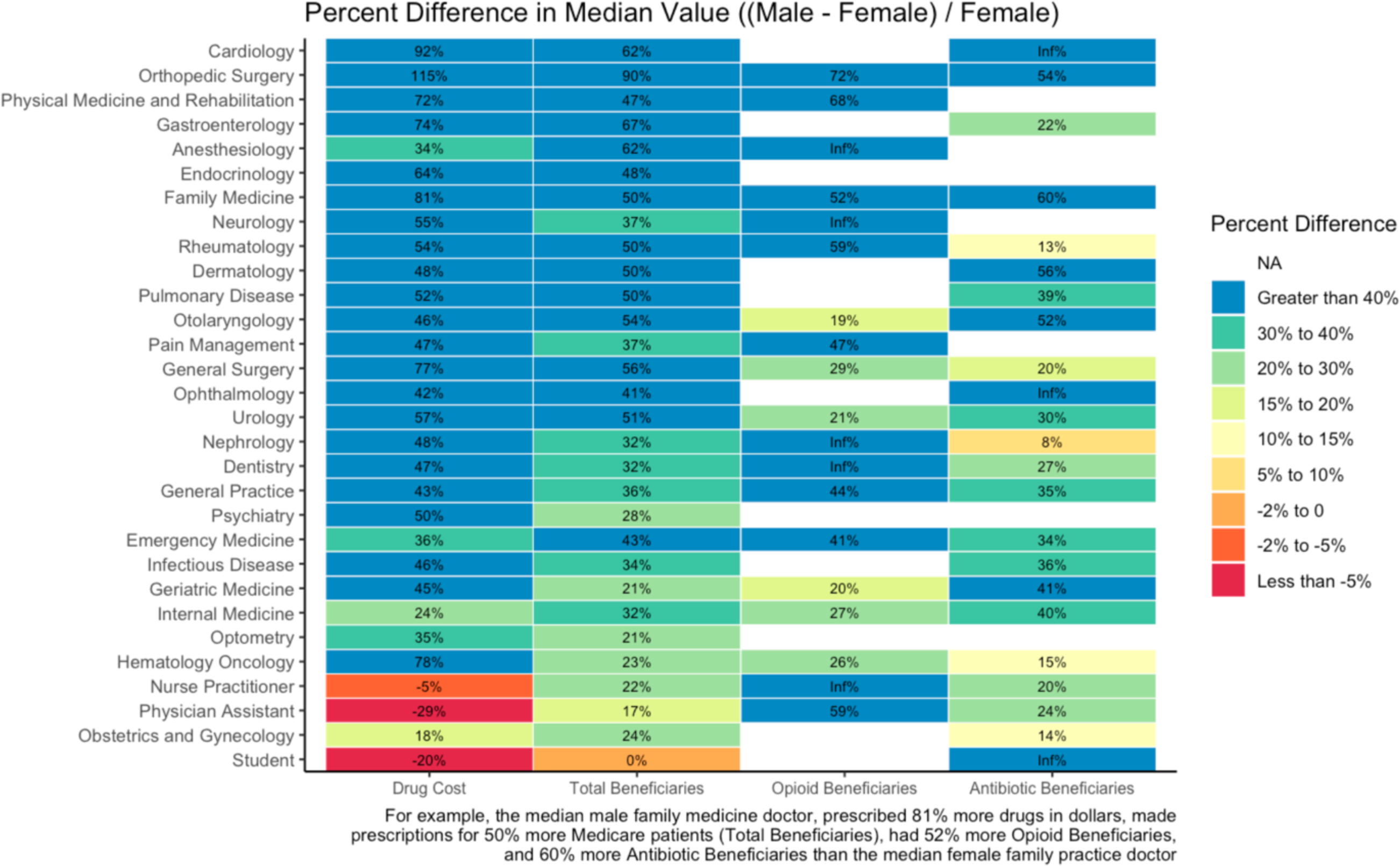
Percent difference of male and female providers’ median drug cost, and Medicare Part D beneficiaries.

The distribution of provider prescribing by specialty and gender for different categories of drugs is shown in Figures 3, 4, 5, and 6. The box plots are sorted by the absolute difference between the median male provider and median female provider within each specialty and category. Figure 3 presents the distribution of claims per beneficiary, and we can see that in almost every specialty, the top quartile, median, and bottom quartiles are higher for male providers, demonstrating that male providers across a wide range of specialties prescribe more drugs per patient than their female counterparts, with the largest differences in Geriatric Medicine and Family Medicine. Figure 4 shows the distribution of opioid prescribing rates by specialty and gender, calculated by CMS as the percentage of all claims made by a provider that are opioid claims. Figure 4 shows that the opioid prescribing rate is higher for male providers in almost every specialty, and the differences are biggest in Anesthesiology, Dentistry, Physical Medicine and Rehabilitation and Neurosurgery. Figure 5 shows the distribution of benzodiazepine claims per beneficiary for the top benzodiazepine prescribing specialties; the biggest differences are in Psychiatry, Family Medicine, and Internal Medicine. Finally, Figure 6 shows the distribution of antibiotic claims per beneficiary for the top antibiotic prescribing specialties; the largest distributional differences are in Family Medicine, Dentistry, and General Practice. Within Urology, female providers prescribed more antibiotics per beneficiary than male providers.

**Figure 2:**
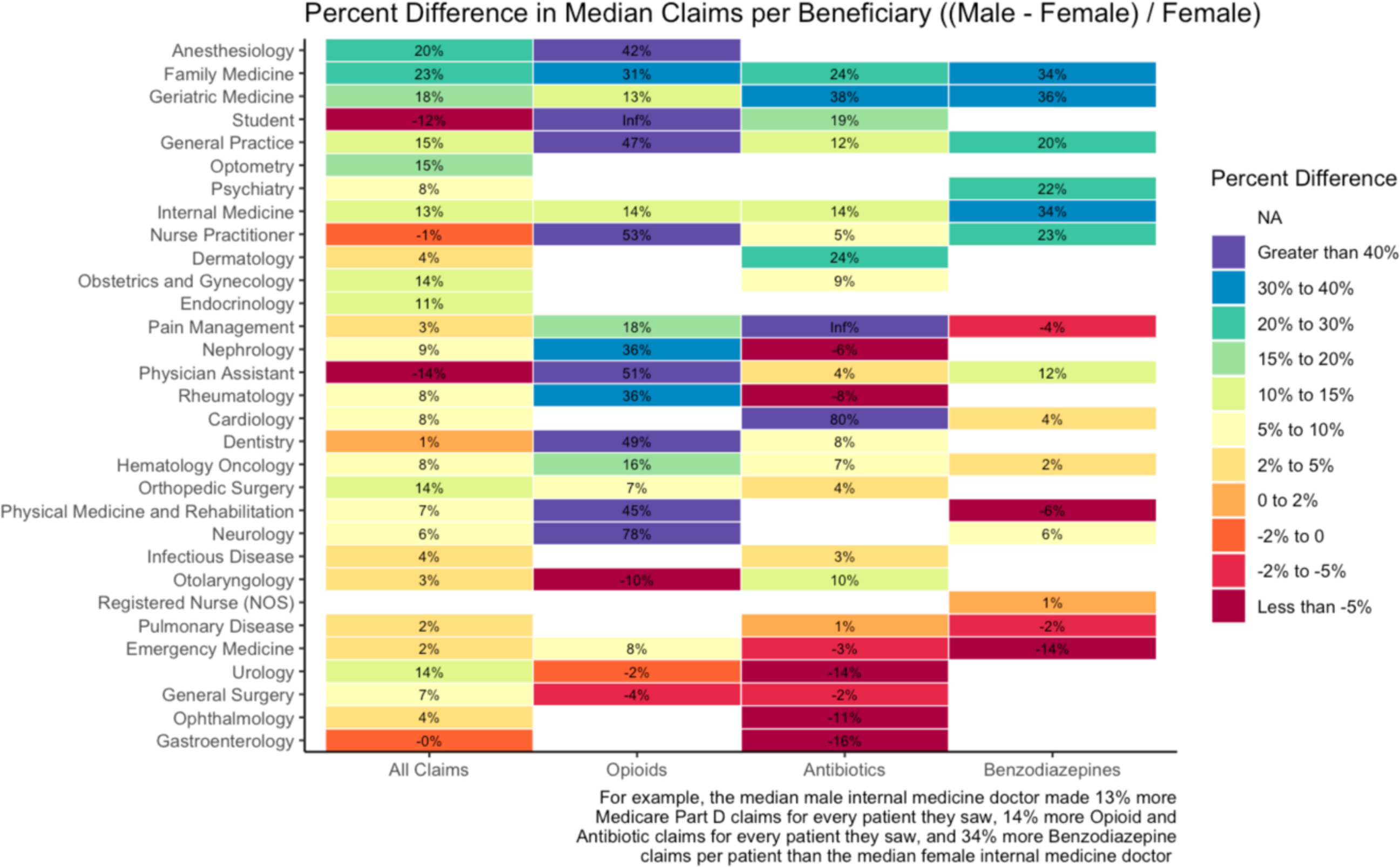
Percent difference of male and female providers’ median drug cost, and Medicare Part D beneficiaries.

**Figure 3:**
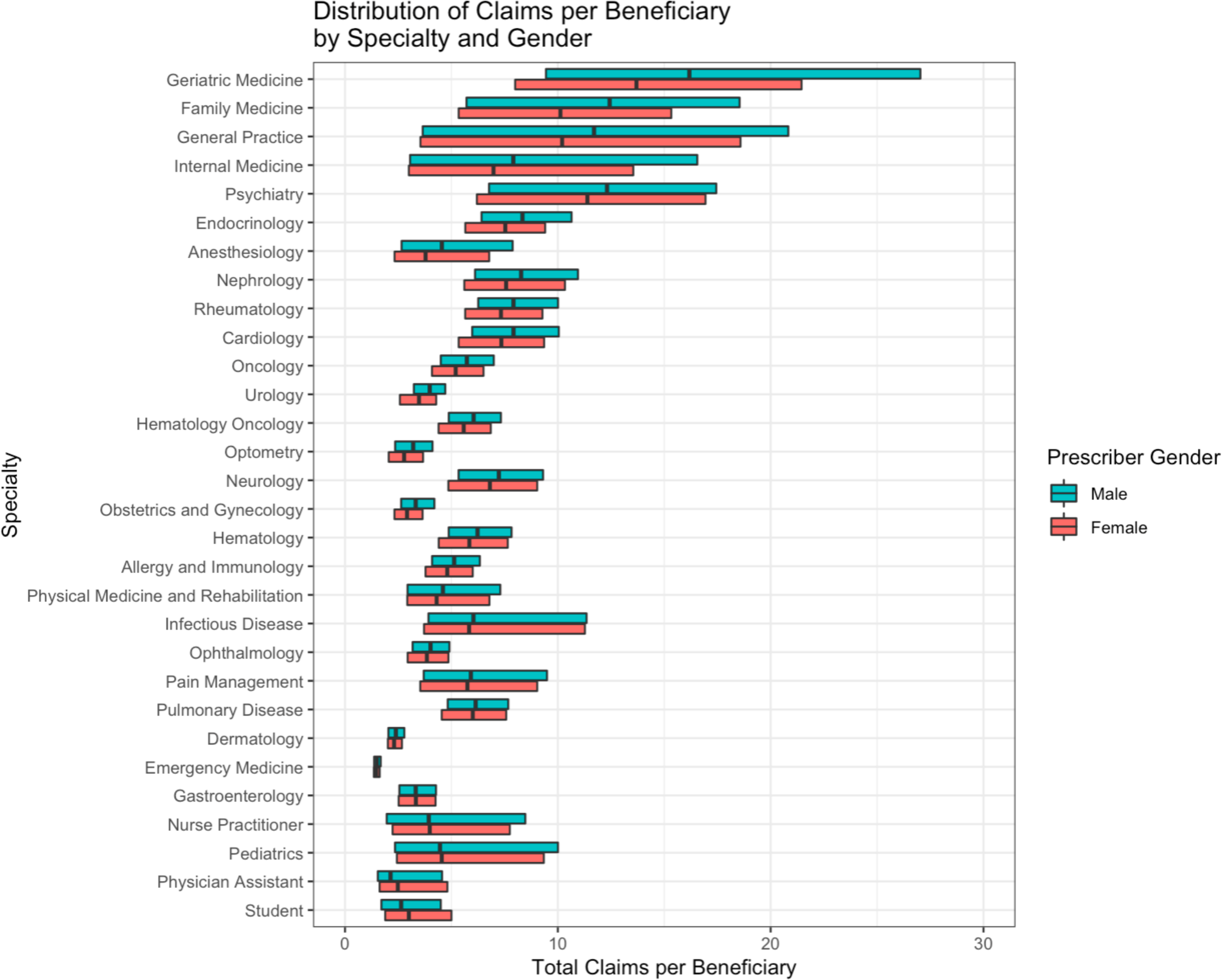
Distribution of provider prescribing of drugs per beneficiary by specialty and gender. From the distribution of claims per beneficiary, it is clear that in almost every specialty, the top quartile, median, and bottom quartiles are higher for male providers, demonstrating that male providers across a wide range of specialties prescribe more drugs per patient than their female counterparts, with the largest differences in Geriatric Medicine and Family Medicine.

**Figure 4:**
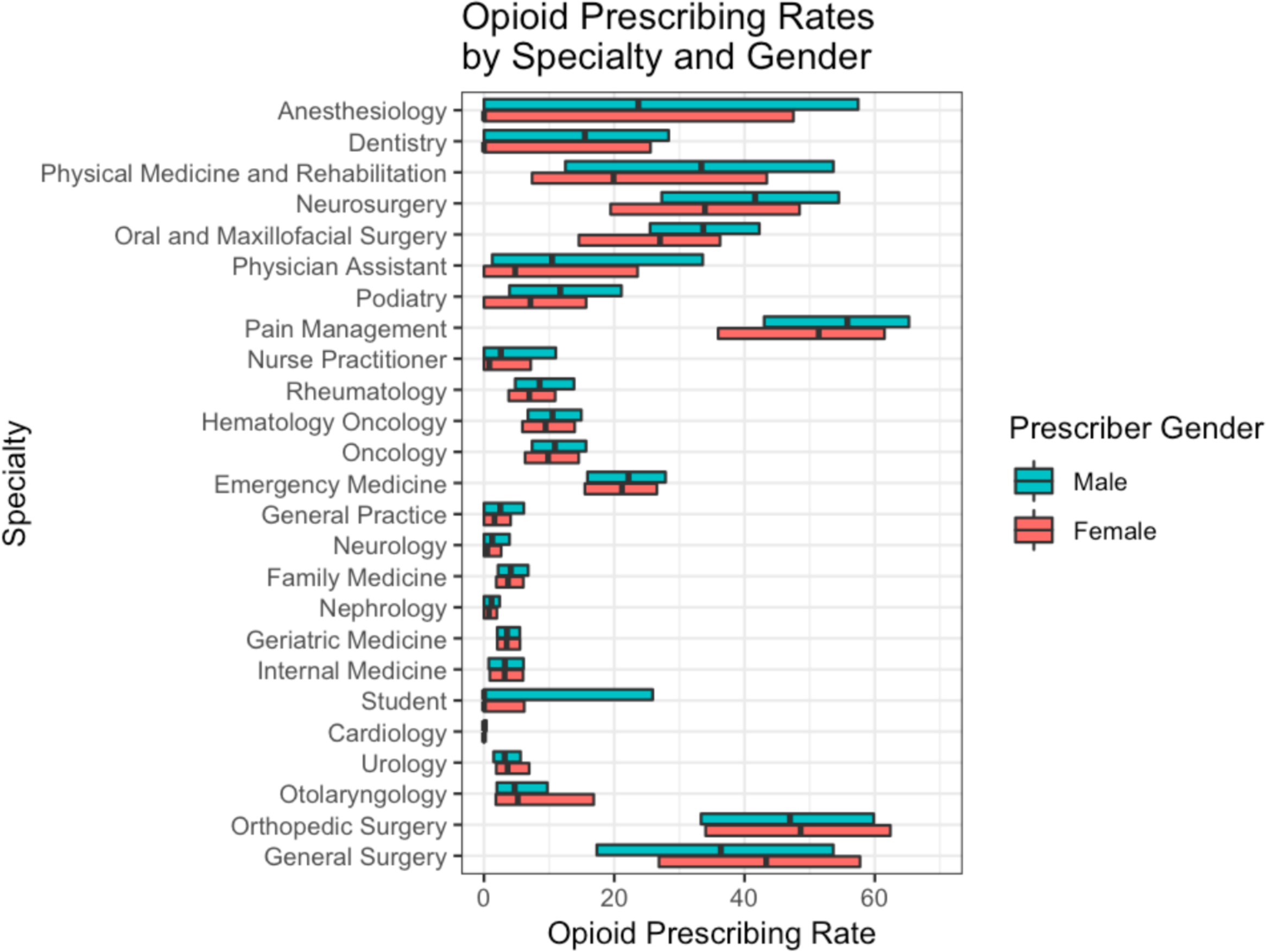
Distribution of provider opioid prescribing rates by specialty and gender. CMS calculates opioid prescribing rates as the percentage of all claims made by a provider that are opioid claims. From the distribution of opioid prescribing rates by specialty and gender, we see that the opioid prescribing rate is higher for male providers in almost every specialty, and the differences are biggest in Anesthesiology, Dentistry, Physical Medicine and Rehabilitation and Neurosurgery.

**Figure 5:**
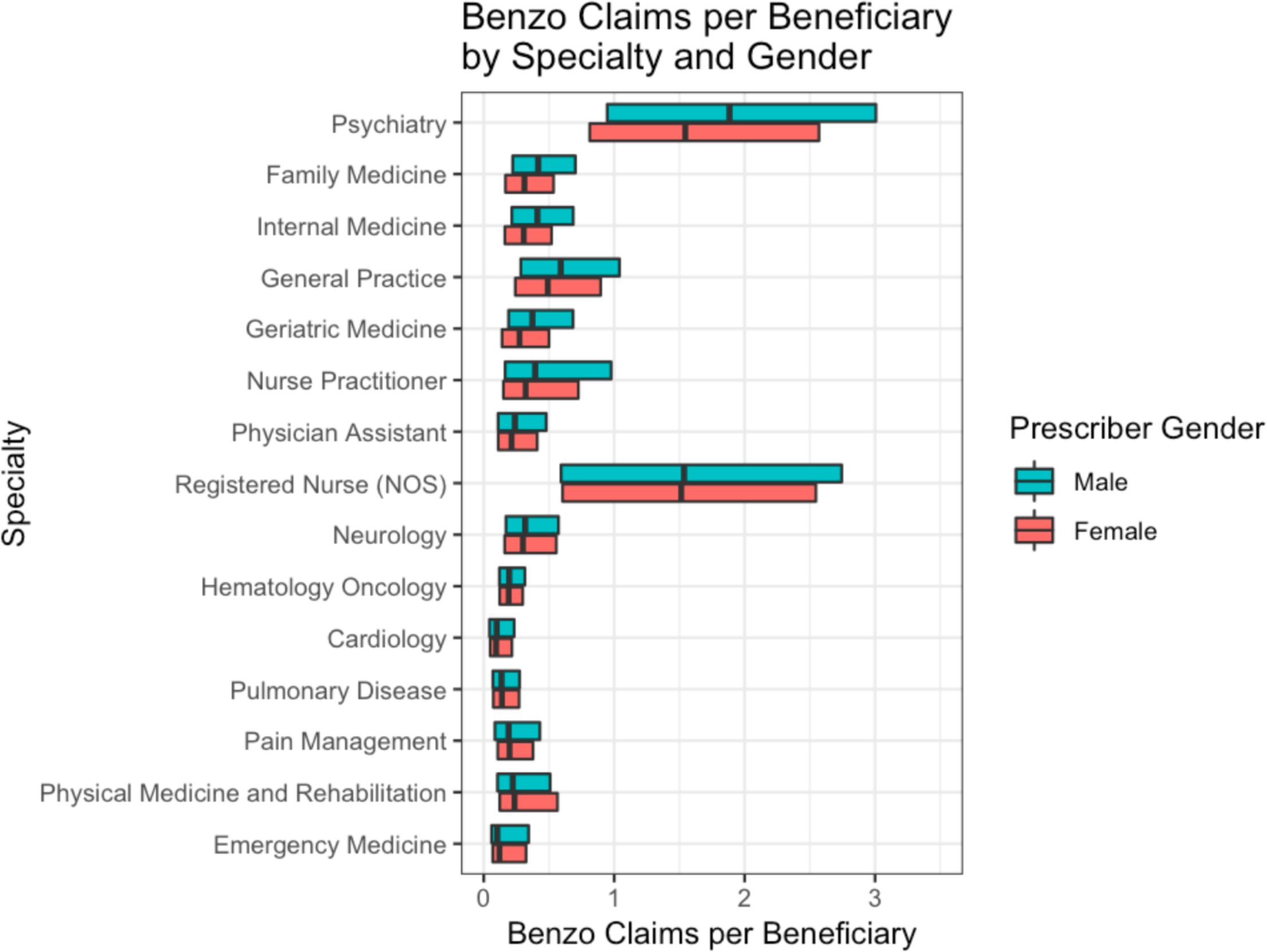
Distribution of provider benzodiazepine prescribing per beneficiary by specialty and gender. The biggest differences in benzodiazepine prescribing are in Psychiatry, Family Medicine, and Internal Medicine.

**Figure 6:**
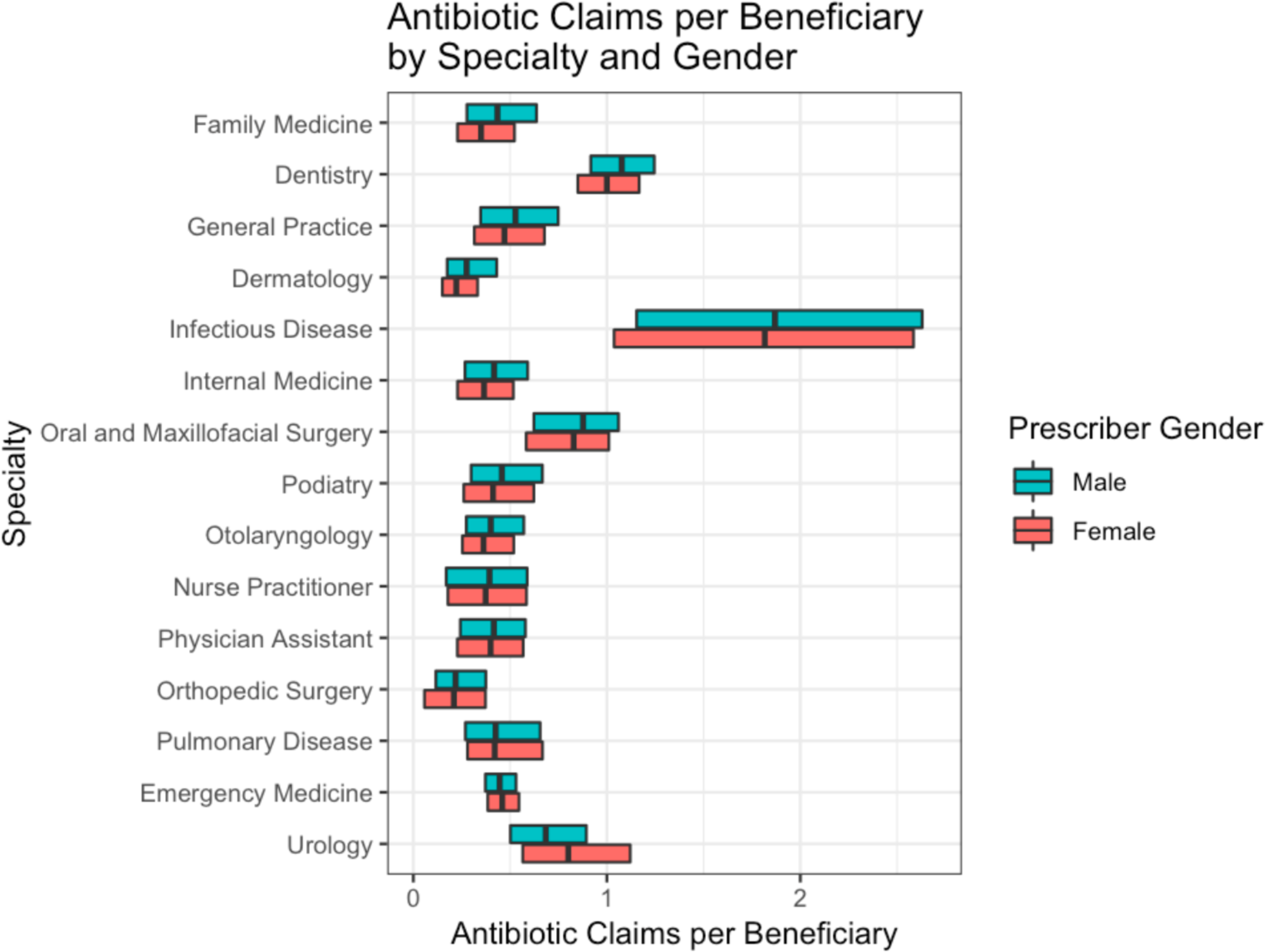
Distribution of provider antibiotic prescribing per beneficiary by specialty and gender. The largest distributional differences are in Family Medicine, Dentistry, and General Practice. Within Urology, female providers prescribed more antibiotics per beneficiary than male providers.

We show the median number of Medicare Part D beneficiaries for male and female providers in different specialties in Figures 7, 8, and 9. The diagonal line in each figure represents parity. Figure 7 shows the median drug beneficiaries. Across every specialty, males had more drug beneficiaries than females. Figure 8 shows the median opioid beneficiaries for every specialty. Again, in every specialty, male providers had more opioid beneficiaries than female providers. Finally, Figure 9 shows the median antibiotic beneficiaries by gender and specialty, and again, male providers had more antibiotic beneficiaries across every specialty.

**Figure 7:**
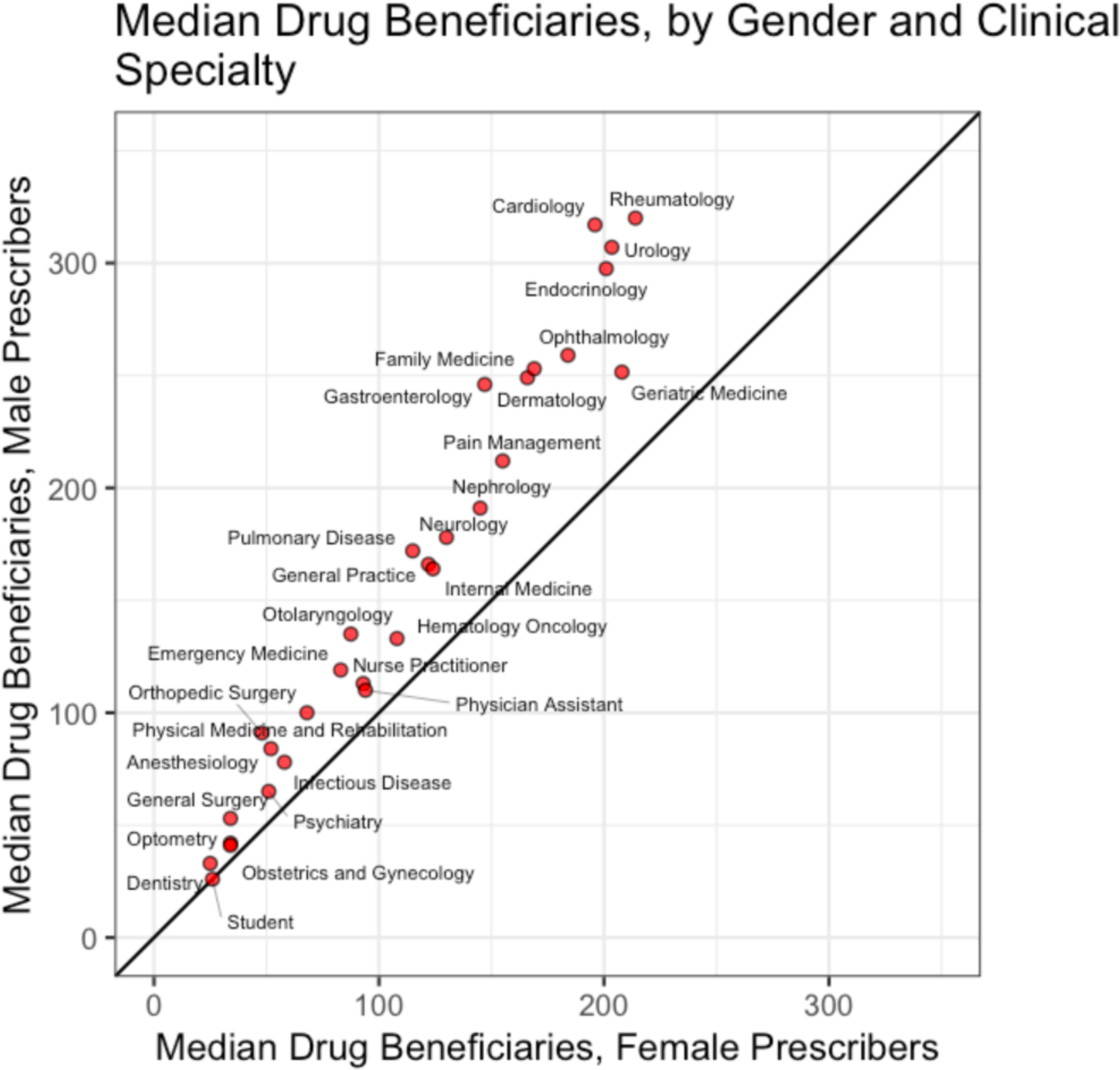
Provider median drug beneficiaries, by gender and specialty. Across every specialty, males had more drug beneficiaries than females.

**Figure 8:**
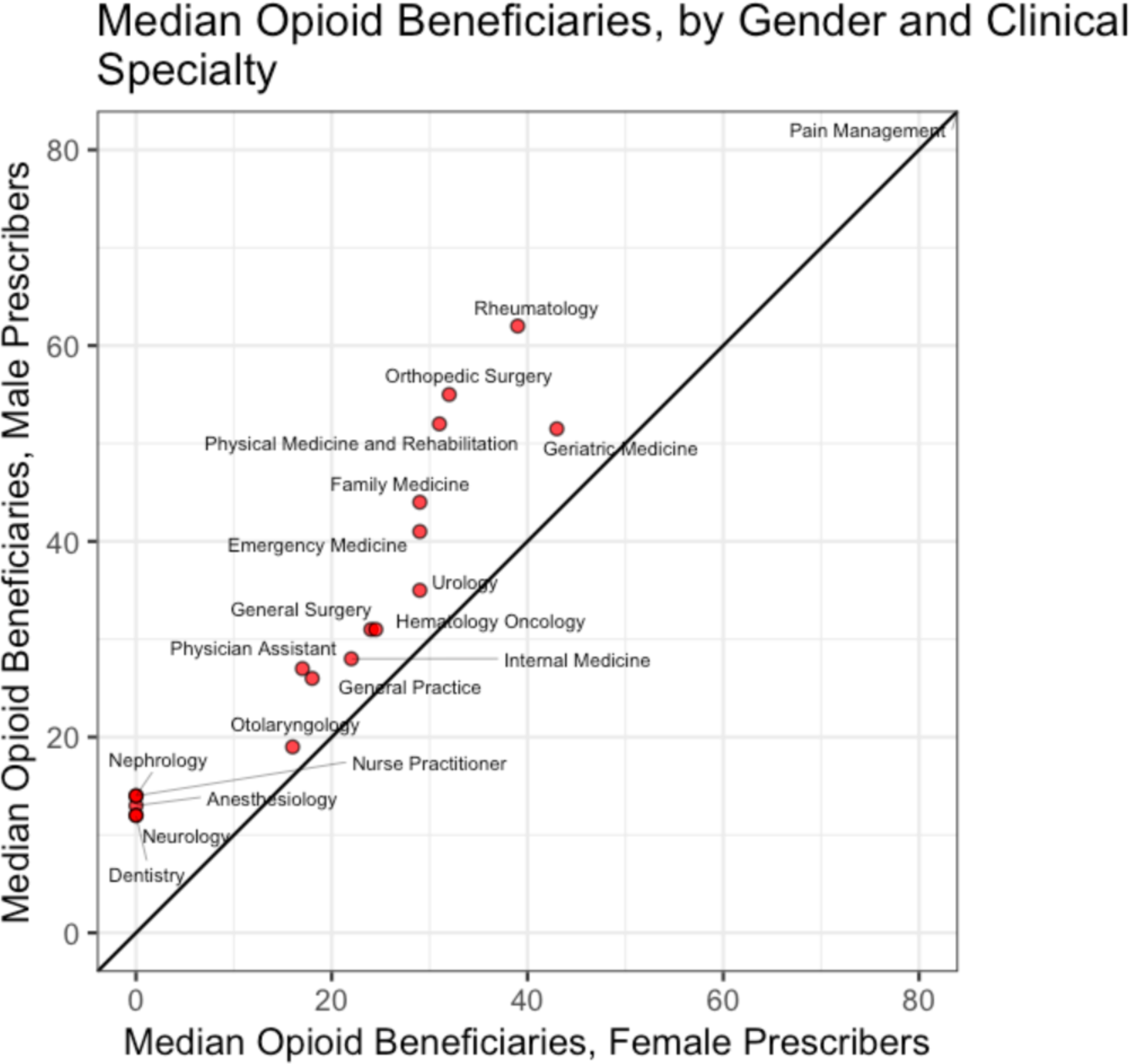
Provider median opioid beneficiaries, by gender and specialty. In every specialty, male providers had more opioid beneficiaries than female providers.

**Figure 9:**
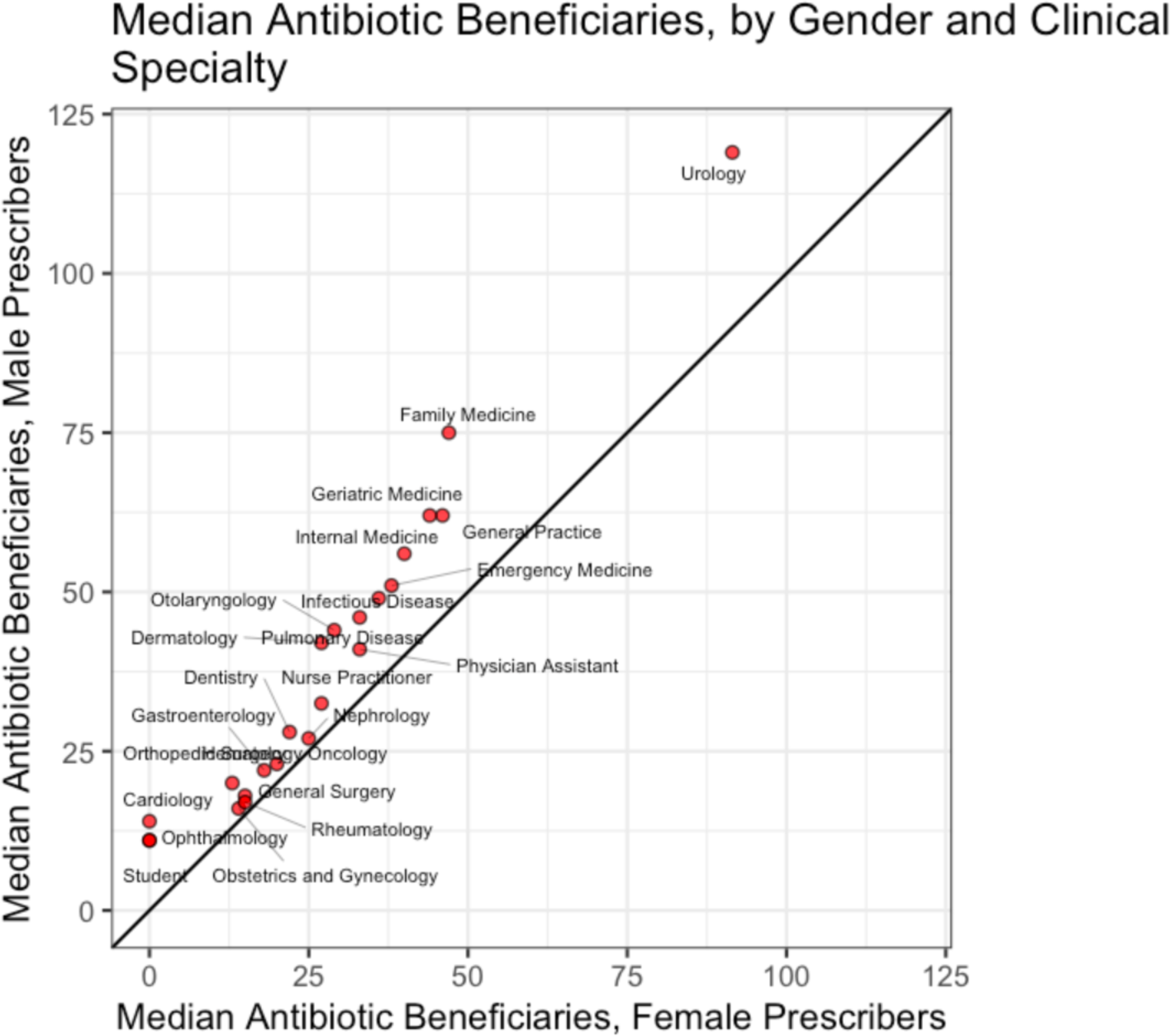
Provider median antibiotic beneficiaries, by gender and specialty. In every specialty, male providers had more antibiotic beneficiaries.

After adjusting for the number of each provider’s patients, the salient trend of male providers making more drug claims persisted. Based on per patient estimates, the heatmap in Figure 2. shows the percent difference of male and female providers’ median total, opioid, benzodiazepine, and antibiotic Medicare Part D drug claims.

After exploring the data visually, we tested the relationships statistically using Mann-Whitney tests. As reported in our table of Mann-Whitney p values (Table 1), in 20 out of 30 top prescribing specialties, male providers made significantly more claims per beneficiary than female providers. In the 15 specialties with the most opioid claims, male providers had significantly higher opioid prescribing rates in 14 specialties (Table 2). In the 15 specialties submitting the most benzodiazepine claims, male providers made significantly more claims per beneficiary than female providers in 7 specialties (Table 3). In the 15 specialties submitting the most antibiotic claims, male providers made significantly more claims per beneficiary than female providers in 9 specialties (Table 4).

**Table 1:** Comparison of the median total claims per beneficiary for male and female providers by specialty.

| <u>Specialty</u> | <u>Median Male</u> | <u>Median Female</u> | <u>Percent Difference</u> | <u>P value</u> | <u>Female Provider Count</u> | <u>Male Provider Count</u> |
| --- | --- | --- | --- | --- | --- | --- |
| Allergy and Immunology | 5.128 | 4.803 | 6.80% | 2.68E-08 | 1,429 | 2,587 |
| Anesthesiology | 4.55 | 3.784 | 20.20% | 0.00000197 | 1,437 | 6,061 |
| Cardiology | 7.917 | 7.346 | 7.80% | <2e-16 | 3,049 | 22,039 |
| Dermatology | 2.388 | 2.305 | 3.60% | 1.89E-10 | 6,141 | 6,890 |
| Emergency Medicine | 1.5 | 1.469 | 2.10% | <2e-16 | 12,955 | 34,274 |
| Endocrinology | 8.338 | 7.531 | 10.70% | <2e-16 | 2,822 | 2,944 |
| Family Medicine | 12.434 | 10.122 | 22.80% | <2e-16 | 44,277 | 65,737 |
| Gastroenterology | 3.322 | 3.329 | -0.20% | 0.335 | 2,249 | 11,101 |
| General Practice | 11.701 | 10.202 | 14.70% | 0.000418 | 2,878 | 7,293 |
| Geriatric Medicine | 16.18 | 13.695 | 18.10% | 0.000000659 | 966 | 962 |
| Hematology | 6.227 | 5.843 | 6.60% | 0.0411 | 219 | 460 |
| Hematology Oncology | 6.042 | 5.578 | 8.30% | <2e-16 | 2,723 | 5,601 |
| Infectious Disease | 6.034 | 5.83 | 3.50% | 0.0641 | 2,070 | 3,183 |
| Internal Medicine | 7.907 | 6.978 | 13.30% | <2e-16 | 49,269 | 83,905 |
| Nephrology | 8.274 | 7.57 | 9.30% | 4.42E-11 | 2,246 | 6,239 |
| Neurology | 7.229 | 6.807 | 6.20% | 5.62E-11 | 4,189 | 9,468 |
| Nurse Practitioner | 3.934 | 3.982 | -1.20% | 0.984 | 124,571 | 13,243 |
| Obstetrics and Gynecology | 3.32 | 2.913 | 14.00% | <2e-16 | 20,018 | 15,680 |
| Oncology | 5.725 | 5.201 | 10.10% | 8.5E-10 | 987 | 2,039 |
| Ophthalmology | 4.017 | 3.847 | 4.40% | 6.86E-11 | 4,694 | 15,069 |
| Optometry | 3.204 | 2.777 | 15.40% | <2e-16 | 10,916 | 17,654 |
| Pain Management | 5.913 | 5.75 | 2.80% | 0.0855 | 558 | 3,302 |
| Pediatrics | 4.462 | 4.545 | -1.80% | 0.58 | 5,298 | 6,460 |
| Physical Medicine and Rehabilitation | 4.6 | 4.304 | 6.90% | 0.0115 | 2,733 | 5,669 |
| Physician Assistant | 2.139 | 2.482 | -13.80% | 1 | 57,697 | 29,036 |
| Psychiatry | 12.312 | 11.39 | 8.10% | 1.17E-12 | 16,387 | 25,631 |
| Pulmonary Disease | 6.138 | 6.004 | 2.20% | 0.00266 | 1,720 | 7,557 |
| Rheumatology | 7.915 | 7.328 | 8.00% | 2.21E-15 | 1,993 | 2,666 |
| Student | 2.636 | 3 | -12.10% | 1 | 22,993 | 28,622 |
| Urology | 3.98 | 3.476 | 14.50% | <2e-16 | 958 | 9,752 |

**Table 2:** Comparison of the median opioid claims per beneficiary for male and female providers by specialty.

| <u>Specialty</u> | <u>Median Male</u> | <u>Median Female</u> | <u>Percent Difference</u> | <u>P value</u> | <u>Female Provider Count</u> | <u>Male Provider Count</u> |
| --- | --- | --- | --- | --- | --- | --- |
| Anesthesiology | 2.391 | 1.683 | 42.10% | 1.14E-10 | 1,129 | 4,996 |
| Dentistry | 0.339 | 0.228 | 48.70% | <2e-16 | 14,857 | 47,290 |
| Emergency Medicine | 0.35 | 0.325 | 7.70% | <2e-16 | 9,739 | 28,419 |
| Family Medicine | 0.507 | 0.387 | 31.00% | <2e-16 | 36,273 | 57,434 |
| General Practice | 0.411 | 0.279 | 47.30% | <2e-16 | 2,127 | 5,851 |
| General Surgery | 0.75 | 0.781 | -4.00% | 1 | 3,401 | 14,571 |
| Hematology Oncology | 0.633 | 0.546 | 15.90% | <2e-16 | 2,308 | 5,022 |
| Internal Medicine | 0.315 | 0.276 | 14.10% | <2e-16 | 37,007 | 66,062 |
| Neurology | 0.119 | 0.067 | 77.60% | <2e-16 | 2,806 | 6,950 |
| Nurse Practitioner | 0.18 | 0.118 | 52.50% | <2e-16 | 94,833 | 10,237 |
| Orthopedic Surgery | 1.004 | 0.935 | 7.40% | 1.38E-12 | 1,119 | 20,685 |
| Pain Management | 3.348 | 2.825 | 18.50% | 0.000287 | 521 | 3,181 |
| Physical Medicine and Rehabilitation | 1.331 | 0.917 | 45.10% | <2e-16 | 2,119 | 4,807 |
| Physician Assistant | 0.37 | 0.245 | 51.00% | <2e-16 | 43,196 | 22,936 |
| Rheumatology | 0.682 | 0.5 | 36.40% | <2e-16 | 1,725 | 2,463 |

**Table 3:** Comparison of the median benzodiazepine claims per beneficiary for male and female providers by specialty.

| **Specialty** | **Median Male** | **Median Female** | **Percent Difference** | **P value** | **Female Provider Count** | **Male Provider Count** |
| --- | --- | --- | --- | --- | --- | --- |
| **Cardiology** | 0.098 | 0.095 | 3.20% | 0.608 | 322 | 4,138 |
| **Emergency Medicine** | 0.103 | 0.12 | -14.20% | 0.99 | 437 | 2,332 |
| **Family Medicine** | 0.417 | 0.311 | 34.10% | <2e-16 | 22,921 | 39,939 |
| **General Practice** | 0.59 | 0.491 | 20.20% | 0.00000108 | 1,212 | 3,479 |
| **Geriatric Medicine** | 0.373 | 0.274 | 36.10% | 7.26E-09 | 627 | 678 |
| **Hematology Oncology** | 0.195 | 0.19 | 2.60% | 0.154 | 1,153 | 2,918 |
| **Internal Medicine** | 0.411 | 0.306 | 34.30% | <2e-16 | 18,116 | 34,781 |
| **Neurology** | 0.318 | 0.3 | 6.00% | 0.0283 | 2,308 | 5,962 |
| **Nurse Practitioner** | 0.394 | 0.321 | 22.70% | <2e-16 | 31,142 | 3,639 |
| **Pain Management** | 0.19 | 0.198 | -4.00% | 0.848 | 202 | 1,554 |
| **Physical Medicine and Rehabilitation** | 0.222 | 0.236 | -5.90% | 0.98 | 627 | 1,861 |
| **Physician Assistant** | 0.239 | 0.213 | 12.20% | 0.00000127 | 9,502 | 4,872 |
| **Psychiatry** | 1.884 | 1.545 | 21.90% | <2e-16 | 8,663 | 15,148 |
| **Pulmonary Disease** | 0.136 | 0.139 | -2.20% | 0.655 | 307 | 2,452 |

**Table 4:** Comparison of the median antibiotic claims per beneficiary for male and female providers by specialty.

| **<u>Specialty</u>** | **<u>Median Male</u>** | **<u>Median Female</u>** | **<u>Percent Difference</u>** | **<u>P value</u>** | **<u>Female Provider Count</u>** | **<u>Male Provider Count</u>** |
| --- | --- | --- | --- | --- | --- | --- |
| **Dentistry** | 1.077 | 1 | 7.70% | <2e-16 | 14,857 | 47,290 |
| **Dermatology** | 0.275 | 0.222 | 23.90% | <2e-16 | 3,984 | 4,171 |
| **Emergency Medicine** | 0.446 | 0.459 | -2.80% | 1 | 9,739 | 28,419 |
| **Family Medicine** | 0.434 | 0.35 | 24.00% | <2e-16 | 36,273 | 57,434 |
| **General Practice** | 0.528 | 0.47 | 12.30% | 3.69E-12 | 2,127 | 5,851 |
| **Infectious Disease** | 1.869 | 1.816 | 2.90% | 0.0294 | 1,441 | 2,139 |
| **Internal Medicine** | 0.417 | 0.365 | 14.20% | <2e-16 | 37,007 | 66,062 |
| **Nurse Practitioner** | 0.393 | 0.373 | 5.40% | 0.0944 | 94,833 | 10,237 |
| **Oral and Maxillofacial Surgery** | 0.877 | 0.828 | 5.90% | 0.000937 | 530 | 5,782 |
| **Orthopedic Surgery** | 0.218 | 0.209 | 4.30% | 0.00443 | 1,119 | 20,685 |
| **Otolaryngology** | 0.4 | 0.362 | 10.50% | 3.85E-09 | 946 | 5,827 |
| **Physician Assistant** | 0.416 | 0.399 | 4.30% | 1.43E-09 | 43,196 | 22,936 |
| **Podiatry** | 0.457 | 0.412 | 10.90% | 2.46E-12 | 2,207 | 8,050 |
| **Pulmonary Disease** | 0.425 | 0.42 | 1.20% | 0.856 | 940 | 4,142 |
| **Urology** | 0.684 | 0.801 | -14.60% | 1 | 714 | 8,051 |

Adjusting for differences in the patient populations of male and female providers, the effect size of male gender on a provider’s overall prescribing rate was significantly positive for 27 out of the top 40 prescribing specialties (Table 5), and significantly negative for 3 specialties. The effect size of male gender on a provider’s opioid prescribing rate was significantly positive for 32 out of 40 specialties, and significantly negative for 1 specialty. The effect size of male gender on a provider’s antibiotic prescribing rate was significantly positive for 20 out of 40 specialties, and significantly negative for 5 specialties. Examples of very larges effect sizes were opioid prescribing in anesthesiology (males made .521 more opioid claims per beneficiary), and overall prescribing in internal medicine (males made 2.268 more claims per beneficiary).

**Table 5:** Effect size of male gender on a provider’s overall prescribing rate, by specialty.

| Specialty | Effect Size Claims per Beneficiary | Effect Size Opioid Claims per Beneficiary | Effect Size Antibiotic Claims per Beneficiary | N (All) | N (Opioids) | N (Antibiotics) |
| --- | --- | --- | --- | --- | --- | --- |
| Allergy and Immunology | 0.294***<br>(0.100, 0.487) | 0.012<br>(-0.012, 0.037) | 0.016<br>(-0.009, 0.041) | 3069 | 2419 | 2348 |
| Anesthesiology | 0.465*<br>(-0.069, 0.999) | 0.521***<br>(0.210, 0.831) | 0.015<br>(-0.015, 0.044) | 3005 | 2847 | 1447 |
| Cardiology | 1.422***<br>(1.266, 1.579) | 0.063***<br>(0.050, 0.075) | 0.031***<br>(0.021, 0.040) | 23398 | 12571 | 10523 |
| Dentistry | 0.042***<br>(0.032, 0.053) | 0.077***<br>(0.070, 0.084) | 0.048***<br>(0.042, 0.053) | 65681 | 37683 | 65274 |
| Dermatology | 0.322***<br>(0.288, 0.356) | -0.033***<br>(-0.041, -0.025) | -0.022***<br>(-0.032, -0.012) | 11780 | 7126 | 10445 |
| Emergency Medicine | 0.190***<br>(0.119, 0.261) | 0.032***<br>(0.023, 0.040) | -0.008***<br>(-0.011, -0.004) | 40557 | 36149 | 39410 |
| Endocrinology | 0.836***<br>(0.632, 1.039) | 0.034***<br>(0.025, 0.044) | 0.033***<br>(0.024, 0.042) | 5235 | 3609 | 2553 |
| Family Medicine | 1.826***<br>(1.677, 1.974) | 0.221***<br>(0.206, 0.235) | 0.127***<br>(0.122, 0.133) | 95798 | 84635 | 88620 |
| Gastroenterology | 0.284***<br>(0.180, 0.389) | 0.025***<br>(0.013, 0.037) | -0.029***<br>(-0.038, -0.020) | 12570 | 6553 | 10636 |
| General Practice | -0.33<br>(-1.000, 0.341) | 0.280***<br>(0.203, 0.358) | 0.064***<br>(0.044, 0.083) | 7462 | 6080 | 6854 |
| General Surgery | 0.533***<br>(0.368, 0.698) | -0.013<br>(-0.035, 0.008) | -0.025**<br>(-0.044, -0.006) | 14070 | 13058 | 10427 |
| Geriatric Medicine | 5.284***<br>(3.853, 6.716) | 0.195***<br>(0.122, 0.269) | 0.138***<br>(0.095, 0.181) | 1749 | 1630 | 1599 |
| Hematology | 0.538*<br>(-0.026, 1.101) | 0.228*<br>(-0.011, 0.467) | -0.016<br>(-0.092, 0.061) | 508 | 401 | 387 |
| Hematology Oncology | 0.328***<br>(0.180, 0.475) | 0.108***<br>(0.080, 0.137) | -0.034***<br>(-0.047, -0.021) | 7110 | 6689 | 5861 |
| Infectious Disease | 0.142<br>(-0.302, 0.585) | 0.050**<br>(0.010, 0.090) | 0.041<br>(-0.023, 0.106) | 3930 | 2541 | 3861 |
| Intensivist | 0.317<br>(-0.407, 1.041) | 0.042<br>(-0.109, 0.192) | 0.052<br>(-0.040, 0.144) | 1070 | 608 | 821 |
| Internal Medicine | 2.268***<br>(2.138, 2.398) | 0.183***<br>(0.172, 0.195) | 0.074***<br>(0.069, 0.079) | 1E+05 | 85622 | 92906 |
| Nephrology | 0.648***<br>(0.398, 0.897) | 0.070***<br>(0.049, 0.092) | -0.01<br>(-0.025, 0.005) | 7818 | 4915 | 6258 |
| Neurology | 0.471***<br>(0.333, 0.609) | 0.224***<br>(0.174, 0.275) | 0.005*<br>(-0.0005, 0.010) | 11391 | 8029 | 6162 |
| Nurse Practitioner | -0.073<br>(-0.203, 0.057) | 0.068***<br>(0.046, 0.091) | 0.019***<br>(0.012, 0.026) | 1E+05 | 80054 | 81249 |
| Obstetrics and Gynecology | 0.380***<br>(0.333, 0.427) | 0.052***<br>(0.032, 0.072) | 0.025***<br>(0.014, 0.036) | 19314 | 7530 | 8037 |
| Oncology | 0.367***<br>(0.147, 0.588) | 0.081***<br>(0.035, 0.126) | -0.016<br>(-0.038, 0.007) | 2312 | 2128 | 1797 |
| Ophthalmology | 0.034<br>(-0.015, 0.084) | 0.015***<br>(0.009, 0.020) | -0.001<br>(-0.008, 0.006) | 17389 | 10382 | 8478 |
| Optometry | 0.299***<br>(0.256, 0.343) | 0.001**<br>(0.0001, 0.002) | 0.012***<br>(0.005, 0.019) | 13414 | 12558 | 5426 |
| Orthopedic Surgery | 0.266***<br>(0.178, 0.355) | 0.169***<br>(0.121, 0.217) | -0.011<br>(-0.034, 0.011) | 18636 | 18349 | 11885 |
| Osteopathy | -2.951***<br>(-5.101, -0.801) | 0.628***<br>(0.209, 1.047) | 0.016<br>(-0.062, 0.093) | 386 | 335 | 309 |
| Otolaryngology | 0.114***<br>(0.043, 0.186) | -0.013<br>(-0.031, 0.005) | 0.041***<br>(0.026, 0.056) | 8679 | 5801 | 7901 |
| Pain Management | -0.412<br>(-0.912, 0.087) | 0.236<br>(-0.054, 0.526) | 0.01<br>(-0.009, 0.029) | 3501 | 3449 | 1602 |
| Pediatrics | -0.765<br>(-1.818, 0.289) | 0.0004<br>(-0.109, 0.110) | 0.031<br>(-0.027, 0.089) | 1464 | 1037 | 1098 |
| Pharmacology | -0.056<br>(-0.289, 0.178) | 0.058*<br>(-0.002, 0.118) | 0.011***<br>(0.003, 0.020) | 1620 | 1571 | 1434 |
| Physical Medicine and Rehabilitation | 0.348**<br>(0.059, 0.638) | 0.567***<br>(0.417, 0.717) | -0.003<br>(-0.024, 0.017) | 6068 | 5611 | 3024 |
| Physician Assistant | -0.071*<br>(-0.148, 0.006) | 0.141***<br>(0.124, 0.158) | 0.019***<br>(0.015, 0.024) | 68953 | 55363 | 55122 |
| Podiatry | 0.080***<br>(0.046, 0.113) | 0.120***<br>(0.104, 0.137) | 0.052***<br>(0.039, 0.065) | 12338 | 8622 | 10887 |
| Psychiatry | 0.166<br>(-0.043, 0.374) | 0.065***<br>(0.044, 0.085) | 0.006***<br>(0.004, 0.008) | 22822 | 15342 | 13272 |
| Pulmonary Disease | 0.413***<br>(0.177, 0.648) | 0.085***<br>(0.051, 0.120) | -0.01<br>(-0.040, 0.020) | 8054 | 4196 | 7387 |
| Registered Nurse (NOS) | 1.378**<br>(0.294, 2.462) | 0.169*<br>(-0.024, 0.362) | 0.071***<br>(0.021, 0.121) | 2073 | 1610 | 1488 |
| Rheumatology | 1.105***<br>(0.899, 1.311) | 0.372***<br>(0.320, 0.424) | -0.007<br>(-0.022, 0.008) | 4321 | 3999 | 2493 |
| Specialist (NOS) | -0.334<br>(-1.482, 0.814) | 0.454**<br>(0.059, 0.849) | 0.067*<br>(-0.006, 0.140) | 767 | 562 | 479 |
| Student | -0.311***<br>(-0.398, -0.224) | 0.070***<br>(0.056, 0.083) | 0.032***<br>(0.023, 0.041) | 20389 | 12002 | 9608 |
| Urology | 0.653***<br>(0.503, 0.804) | -0.001<br>(-0.024, 0.023) | 0.155***<br>(0.127, 0.184) | 8843 | 7571 | 8742 |

Covariates included in linear model: provider gender; male ratio of providers' patients; average age of providers' beneficiaries; average risk score of providers' beneficiaries p-values: \*\*\*: .01; \*\*: .05; \*: .1

After adjusting for differences in the patient populations of male and female providers, we sought to control for other provider differences besides gender. We approximated the age of providers by calculating the number of years they have practiced medicine (calculated as 2016 minus the year in which they graduated from medical school). After filtering our data to include only providers for whom we have medical school graduation data, the number of providers in our dataset got reduced by approximately one third. The Pearson correlation of the age and gender of providers is .36 and statistically significant with a p-value of less than 2.2e-16. Table 6 shows the effect sizes of gender on prescribing per patient after adjusting for both the differences in the patient populations of male and female providers as well as for the age of providers.

**Table 6:**
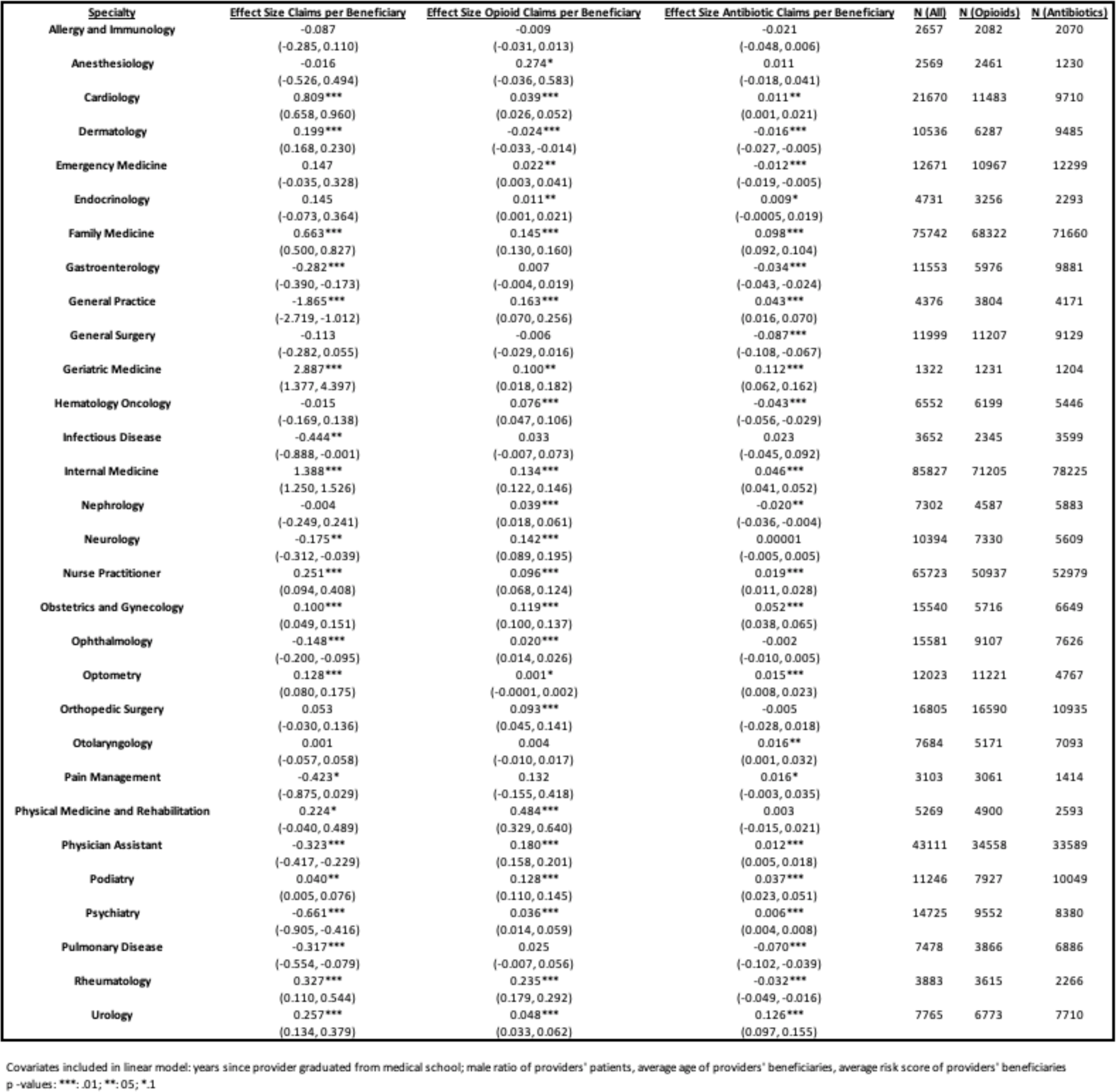
Effect size of male gender on a provider’s overall prescribing rate (including additional covariates), by specialty.

| Specialty | Effect Size Claims per Beneficiary | Effect Size Opioid Claims per Beneficiary | Effect Size Antibiotic Claims per Beneficiary | N (All) | N (Opioids) | N (Antibiotics) |
| --- | --- | --- | --- | --- | --- | --- |
| Allergy and Immunology | -0.087<br>(-0.285, 0.110) | -0.009<br>(-0.031, 0.013) | -0.021<br>(-0.048, 0.006) | 2657 | 2082 | 2070 |
| Anesthesiology | -0.016<br>(-0.526, 0.494) | 0.274*<br>(-0.036, 0.583) | 0.011<br>(-0.018, 0.041) | 2569 | 2461 | 1230 |
| Cardiology | 0.809***<br>(0.658, 0.960) | 0.039***<br>(0.026, 0.052) | 0.011**<br>(0.001, 0.021) | 21670 | 11483 | 9710 |
| Dermatology | 0.199***<br>(0.168, 0.230) | -0.024***<br>(-0.033, -0.014) | -0.016***<br>(-0.027, -0.005) | 10536 | 6287 | 9485 |
| Emergency Medicine | 0.147<br>(-0.035, 0.328) | 0.022**<br>(0.003, 0.041) | -0.012***<br>(-0.019, -0.005) | 12671 | 10967 | 12299 |
| Endocrinology | 0.145<br>(-0.073, 0.364) | 0.011**<br>(0.001, 0.021) | 0.009*<br>(-0.0005, 0.019) | 4731 | 3256 | 2293 |
| Family Medicine | 0.663***<br>(0.500, 0.827) | 0.145***<br>(0.130, 0.160) | 0.098***<br>(0.092, 0.104) | 75742 | 68322 | 71660 |
| Gastroenterology | -0.282***<br>(-0.390, -0.173) | 0.007<br>(-0.004, 0.019) | -0.034***<br>(-0.043, -0.024) | 11553 | 5976 | 9881 |
| General Practice | -1.865***<br>(-2.719, -1.012) | 0.163***<br>(0.070, 0.256) | 0.043***<br>(0.016, 0.070) | 4376 | 3804 | 4171 |
| General Surgery | -0.113<br>(-0.282, 0.055) | -0.006<br>(-0.029, 0.016) | -0.087***<br>(-0.108, -0.067) | 11999 | 11207 | 9129 |
| Geriatric Medicine | 2.887***<br>(1.377, 4.397) | 0.100**<br>(0.018, 0.182) | 0.112***<br>(0.062, 0.162) | 1322 | 1231 | 1204 |
| Hematology Oncology | -0.015<br>(-0.169, 0.138) | 0.076***<br>(0.047, 0.106) | -0.043***<br>(-0.056, -0.029) | 6552 | 6199 | 5446 |
| Infectious Disease | -0.444**<br>(-0.888, -0.001) | 0.033<br>(-0.007, 0.073) | 0.023<br>(-0.045, 0.092) | 3652 | 2345 | 3599 |
| Internal Medicine | 1.388***<br>(1.250, 1.526) | 0.134***<br>(0.122, 0.146) | 0.046***<br>(0.041, 0.052) | 85827 | 71205 | 78225 |
| Nephrology | -0.004<br>(-0.249, 0.241) | 0.039***<br>(0.018, 0.061) | -0.020**<br>(-0.036, -0.004) | 7302 | 4587 | 5883 |
| Neurology | -0.175**<br>(-0.312, -0.039) | 0.142***<br>(0.089, 0.195) | 0.00001<br>(-0.005, 0.005) | 10394 | 7330 | 5609 |
| Nurse Practitioner | 0.251***<br>(0.094, 0.408) | 0.096***<br>(0.068, 0.124) | 0.019***<br>(0.011, 0.028) | 65723 | 50937 | 52979 |
| Obstetrics and Gynecology | 0.100***<br>(0.049, 0.151) | 0.119***<br>(0.100, 0.137) | 0.052***<br>(0.038, 0.065) | 15540 | 5716 | 6649 |
| Ophthalmology | -0.148***<br>(-0.200, -0.095) | 0.020***<br>(0.014, 0.026) | -0.002<br>(-0.010, 0.005) | 15581 | 9107 | 7626 |
| Optometry | 0.128***<br>(0.080, 0.175) | 0.001*<br>(-0.0001, 0.002) | 0.015***<br>(0.008, 0.023) | 12023 | 11221 | 4767 |
| Orthopedic Surgery | 0.053<br>(-0.030, 0.136) | 0.093***<br>(0.045, 0.141) | -0.005<br>(-0.028, 0.018) | 16805 | 16590 | 10935 |
| Otolaryngology | 0.001<br>(-0.057, 0.058) | 0.004<br>(-0.010, 0.017) | 0.016**<br>(0.001, 0.032) | 7684 | 5171 | 7093 |
| Pain Management | -0.423*<br>(-0.875, 0.029) | 0.132<br>(-0.155, 0.418) | 0.016*<br>(-0.003, 0.035) | 3103 | 3061 | 1414 |
| Physical Medicine and Rehabilitation | 0.224*<br>(-0.040, 0.489) | 0.484***<br>(0.329, 0.640) | 0.003<br>(-0.015, 0.021) | 5269 | 4900 | 2593 |
| Physician Assistant | -0.323***<br>(-0.417, -0.229) | 0.180***<br>(0.158, 0.201) | 0.012***<br>(0.005, 0.018) | 43111 | 34558 | 33589 |
| Podiatry | 0.040**<br>(0.005, 0.076) | 0.128***<br>(0.110, 0.145) | 0.037***<br>(0.023, 0.051) | 11246 | 7927 | 10049 |
| Psychiatry | -0.661***<br>(-0.905, -0.416) | 0.036***<br>(0.014, 0.059) | 0.006***<br>(0.004, 0.008) | 14725 | 9552 | 8380 |
| Pulmonary Disease | -0.317***<br>(-0.554, -0.079) | 0.025<br>(-0.007, 0.056) | -0.070***<br>(-0.102, -0.039) | 7478 | 3866 | 6886 |
| Rheumatology | 0.327***<br>(0.110, 0.544) | 0.235***<br>(0.179, 0.292) | -0.032***<br>(-0.049, -0.016) | 3883 | 3615 | 2266 |
| Urology | 0.257***<br>(0.134, 0.379) | 0.048***<br>(0.033, 0.062) | 0.126***<br>(0.097, 0.155) | 7765 | 6773 | 7710 |

Covariates included in linear model: years since provider graduated from medical school; male ratio of providers' patients, average age of providers' beneficiaries, average risk score of providers' beneficiaries p-values: \*\*\*: .01; \*\*: .05; \*: .1

As Table 6 shows, the effect size of male gender on a provider’s overall prescribing rate was significantly positive for 12 out of the top 30 prescribing specialties, and significantly negative for 9 specialties; the effect size of male gender on a provider’s opioid prescribing rate was significantly positive for 22 out of 30 specialties, and significantly negative for 1 specialty; and the effect size of male gender on a provider’s antibiotic prescribing rate was significantly positive for 15 out of 30 specialties, and significantly negative for 8 specialties. Examples of very larges effect sizes were opioid prescribing in anesthesiology (males made .274 more opioid claims per beneficiary), and overall prescribing in internal medicine (males made 1.388 more claims per beneficiary).

## Discussion

Our population-based study of Medicare Part D patients found that male providers prescribed more drugs by volume and cost, as well as per patient, including more opioids, benzodiazepines, and antibiotics, than female providers. The etiology of these observed gender differences in prescribing is unknown, and is likely to be complex and multifactorial. Areas of future research could include the potential impact of patient volume, pharmaceutical payments, and/or gendered cultural differences.

Our data imply that male prescribers see more Medicare patients in a year, although we were not able to assess whether patient volume in fact reduces time per patient. Nonetheless, systemic and financial pressures to see more patients per unit time, and have ‘satisfied customers,’ may contribute to increased prescribing.^5^ Previous research has found that male physicians spend less time with their patients than female physicians^6^: shorter visits have been associated with increased prescribing.^7^ Female providers are more likely to engage in prevention-rich treatments than their male counterparts.^8,9,10,11,12^ Male physicians are more likely than females to receive pharmaceutical payments, which has been linked to higher prescribing of the marketed drug,^13^ and may also play a role in these data. Our data found the starkest gender differences in opioid prescribing in specialties that target pain. A study of opioid prescribing to chronic pain patients in France found no gender differences,^14^ suggesting the intersection between culture, opioids, and pain, may lead to gendered differences in prescribing.

Our analysis had the following key limitations. Our dataset did not necessarily reflect clinicians’ complete practice, and did not allow us to adjust for individual patient-level variables such as medical complexity. In some specialties, there was either a small sample size of female providers (urology), or a large gender imbalance (orthopedic surgery). We demonstrate significant findings, based on administrative drug claims for Medicare beneficiaries; their generalization to the population at-large is unknown.

## Data Availability

The data used for this study is publicly available and hosted by CMS.

https://www.cms.gov/Research-Statistics-Data-and-Systems/Statistics-Trends-and-Reports/Medicare-Provider-Charge-Data/PartD2016.html

## Conclusion

Male Medicare part D providers across diverse specialties are more likely than female providers to prescribe all types of medications, including opioids, benzodiazepines, and antibiotics. This finding informs the design of prevention strategies that seek to reduce iatrogenic harms related to prescribing. Our study is the first study showing gender differences in prescribing by specialty at the population level. Future research could consider mixed methods approaches that include the analysis of more granular data sources, such as those that can be linked directly to each patient’s comprehensive care record, and qualitative studies looking at the social, economic, and cultural factors driving these observed gender differences.

## Notes

### Competing Interest Statement

The authors have declared no competing interest.

### Funding Statement

No external funding received.

### Author Declarations

All relevant ethical guidelines have been followed and any necessary IRB and/or ethics committee approvals have been obtained.

Any clinical trials involved have been registered with an ICMJE-approved registry such as ClinicalTrials.gov and the trial ID is included in the manuscript.

